# Multi-chain Fudan-CCDC model for COVID-19 – a revisit to Singapore’s case

**DOI:** 10.1101/2020.04.13.20063792

**Authors:** Hanshuang Pan, Nian Shao, Yue Yan, Xinyue Luo, Shufen Wang, Ling Ye, Jin Cheng, Wenbin Chen

**Affiliations:** School of Mathematical Sciences, Fudan University, 220 Handan Road, Shanghai, 200433, China; Shanghai Key Laboratory for Contemporary Applied Mathematics, Fudan University, 220 Handan Road, Shanghai, 200433, China; School of Mathematics, Shanghai University of Finance and Economics, 777 Guoding Road, Shanghai, 200433, China; Daishan County Center for Disease Control and Prevention, 189 Penglai Road, Daishan, Zhejiang, 316200, China

**Keywords:** COVID-19, Singapore, multi-chain Fudan-CCDC model

## Abstract

**Background:** COVID-19 has been impacting on the whole world critically and constantly since late December 2019. Rapidly increasing infections has raised intense world-wide attention. How to model the evolution of COVID-19 effectively and efficiently is of great significance for prevention and control.

**Methods:** We propose the multi-chain Fudan-CCDC model based on the original single-chain model in [8] to describe the evolution of COVID-19 in Singapore. Multi-chains can be considered as the superposition of several single chains with different characteristics. We identify parameters of models by minimizing the penalty function.

**Results:** The numerical simulation results exhibit the multichain model performs well on data fitting. Though unsteady the increments are, they could still fall within the range of ±25% fluctuation from simulation results. It is predicted by multi-chain models that Singapore are experiencing a nonnegligible risk of explosive outbreak, thus stronger measures are urgently needed to contain the epidemic.

**Conclusion:** The multi-chain Fudan-CCDC model provides an effective way to early detect the appearance of imported infectors and super spreaders and forecast a second outbreak. It can also explain the data in those countries where the single-chain model shows deviation from the data.

## 1 Introduction

The Fudan-CCDC model [9, 8, 12] was proposed by Cheng’s group at Fudan University to study the evolution of COVID-19. The model took advantages of the time delay process introduced by the TDD-NCP model [13, 4, 3, 6, 7, 11] proposed previously also by Cheng’s group, and developed new convolution kernels for the time delay terms by applying several time distributions acquired from an important paper [5] by CCDC (China Center for Disease Control and Prevention). Both the TDD-NCP model and the Fudan-CCDC model are single-chain models, and have been performed well in analyzing the evolution of COVID-19 in China, and its early stage of global transmission [10, 14]. Since Mar 2, some of the simulation results on South Korea, Iran, Italy, Germany, Spain, the U.S. et al have been posted on Twitter_@fudan ccdc.

The idea of multi-chain was first introduced by Nian Shao and Wenbin Chen on Feb 19, 2020, when they were tracking the curve of cumulative confirmed cases in South Korea. They found that there was a sudden turn in growth rate, just as described in Figure 1, shown in a semi-log way. Our group suspected that a stronger transmission chain had occurred, so we began to develop a multi-chain Fudan-CCDC model. Based on the new model and the public data, we identified the start date of the new transmission chain in South Korea was around Feb 10, and thus held the view that there was a new source emerging at that time. Then we announced our finding [2] on Feb 20. Later, it was confirmed by the news [1] that there was indeed a case of super spreader on Feb 7.

**Figure 1:**
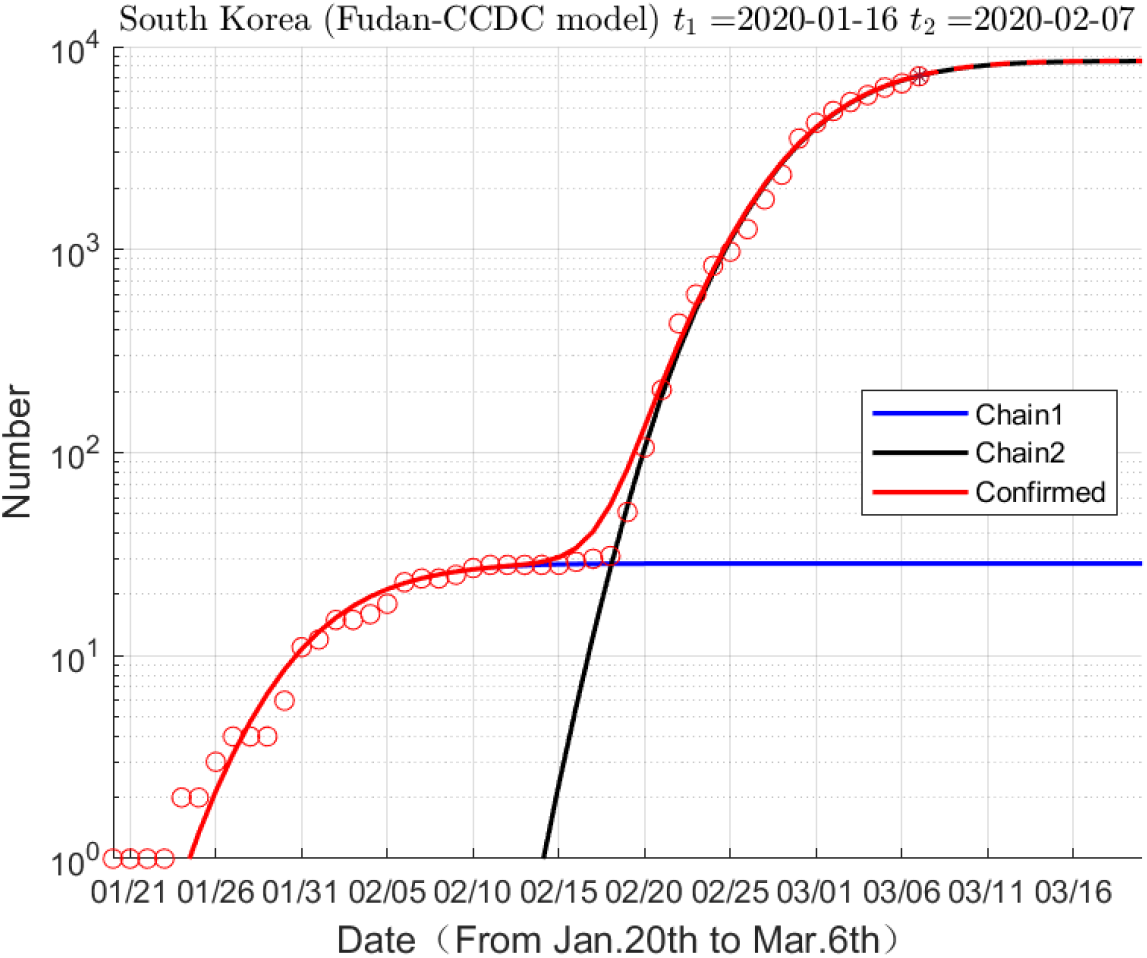
Semi-log plot of data fitting for South Korea, suggesting two chains in transmission. Red circles: data, daily number of cumulative confirmed cases in South Korea, Jan 20 to Mar 6; Red line: curve fitting for data; Blue line: the first chain; Black line: the second chain. The red, blue, black lines are all estimated by the model.

With the further spread of the global pandemic, such sudden change of growth rate has been observed in the cases of other countries as well. And the curves fitted by the Fudan-CCDC model sometimes deviate from the data. Singapore is one of the examples. We had studied Singapore’s case in [14], and based on the data till Feb 25, we concluded that Singapore had been successful in disease prevention and control. Since then, one member of our group, Hanshuang Pan, has been continually tracking the data. Unexpectedly, in late February, a sudden rise occurred, see Figure 2(b).

**Figure 2:**
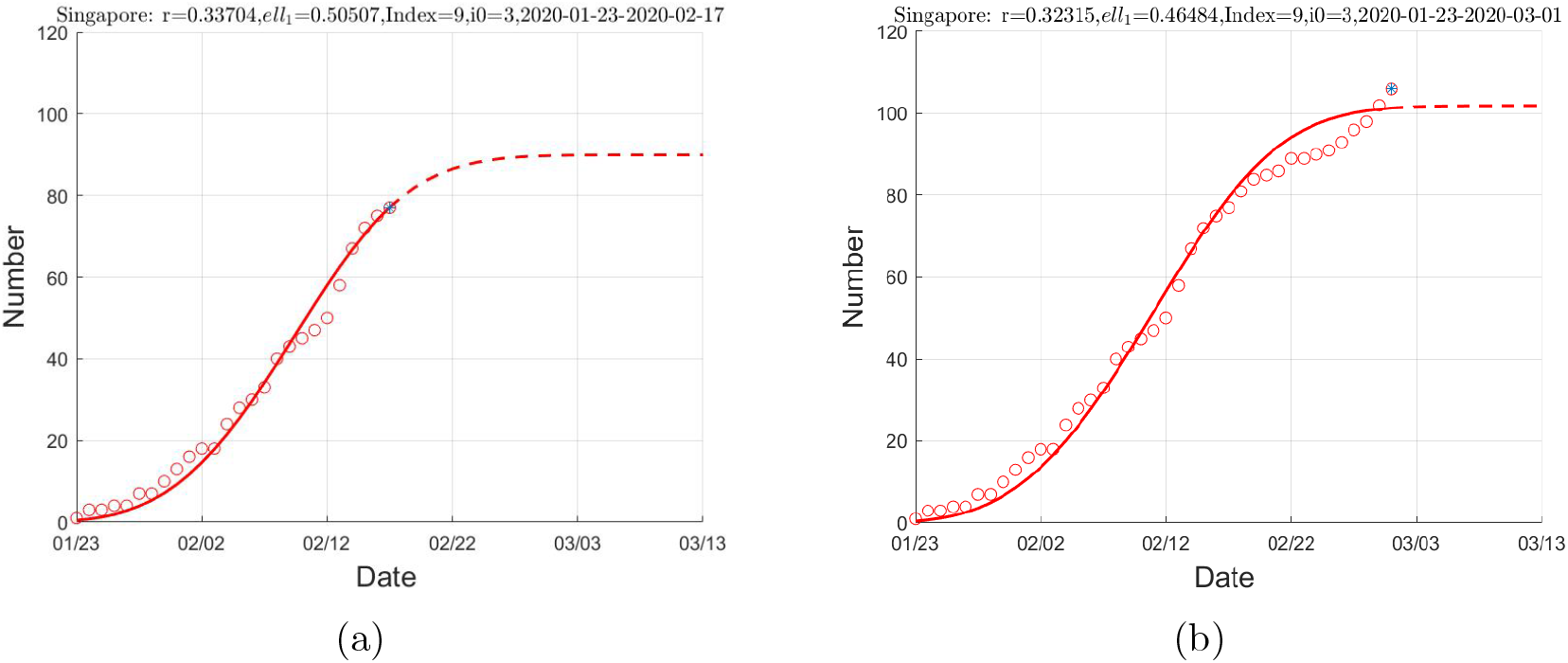
Data fitting for Singapore. (a) Jan 23 to Feb 17; (b) Jan 23 to Mar 1. Red circles: data, daily number of cumulative confirmed cases in Singapore; Red line: curve fitting for data; Dotted line: predictions.

We show in Figure 2 the curve fitting for Singapore’s data by the Fudan-CCDC model, on Feb 17 and Mar 1, respectively. We see in Figure 2(a) that on Feb 17, the Fudan-CCDC model had predicted that the increment of confirmed cases would be zero on Feb 27, and remained stable for the next ten days. However, on the crucial day Feb 27, an unexpected rise occurred in Singapore’s data (see Figure 2(b)), which caused our vigilance. Till Mar 1, this new upward trend was so obvious that it could not be explained by the single chain Fudan-CCDC model any longer. Therefore, we began to consider the application of the multi-chain Fudan-CCDC model, and revisit Singapore’s case.

## 2 Results

Through simulations, we find that the evolution of the epidemic so far can be well explained by the two-chain and the three-chain Fudan-CCDC model, although in real-life transmissions, the number of chains may be far more than three. In this section, we display the results of the two-chain and three-chain models, and based on the data till Apr 12, we warn that Singapore is in the risk of a sudden increase in the number of infected people.

### 2.1 Results of the two-chain model

Figure 3 and Figure 4 show the evolutions of COVID-19 in Singapore and its possible future trends, based on the two-chain model. The scattered red circles are the data: the number of cumulative confirmed cases (Figure 3) and its increment (Figure 4) from Jan 23 to Apr 4. In Figure 3, we illustrate the four ‘most optimized’ fitting curves (in solid lines) for the data, and their predictions (in dotted lines) by the model, in the order of red, green, blue and purple, respectively. For the convenience to recognize, the ‘very most optimized’ fitting curve is drawn in a full solid red line, including its prediction. Details of the optimization methods are described in the section Materials and Methods. We can see from Figure 3(a) that based on the two-chain model, Singapore is expected to have zero increment of confirmed cases on Apr 26, and the total number of infections will be around 1500, if no other transmission chains arise in the future. Figure 3(b) is the semi-log form of Figure 3(a), and it clearly demonstrates the excellent curve fitting of the sudden rise in growth rate around Mar 3.

**Figure 3:**
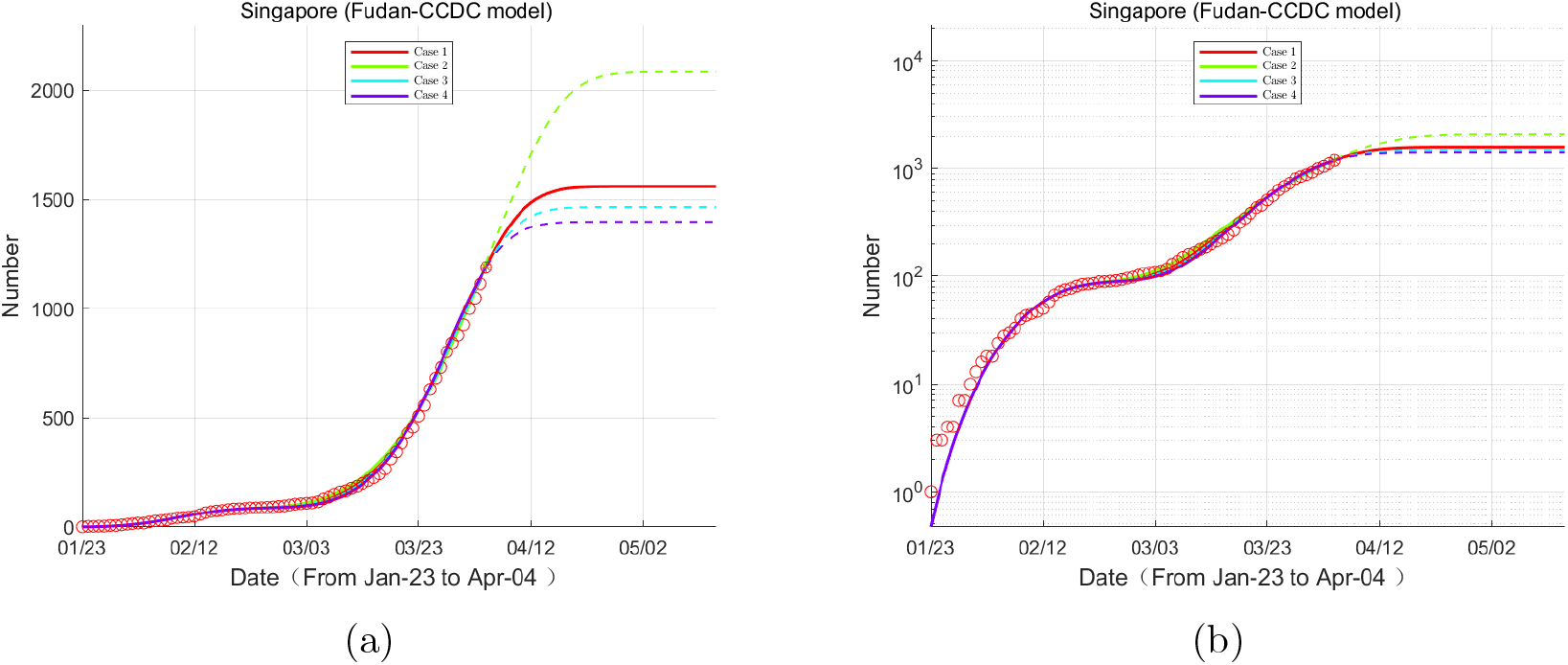
(a) Evolution of number of cumulative confirmed cases based on the two-chain Fudan-CCDC model. Red circles: data; Red, green, blue and purple lines: the four ‘most optimized’ fitting curves and their predictions. (b) The semi-log form of (a).

**Figure 4:**
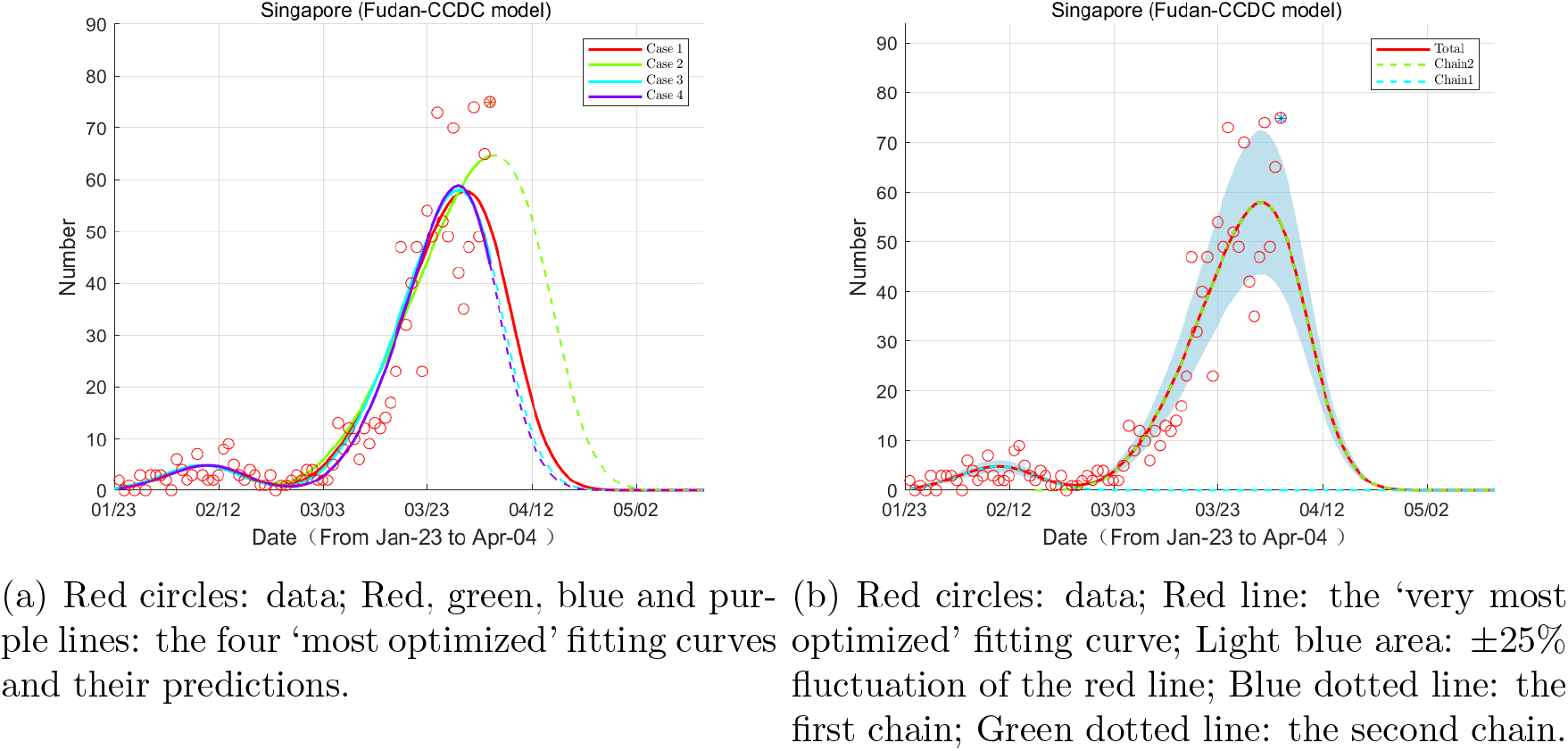
Evolution of the increment of confirmed cases based on the two-chain Fudan-CCDC model.

Figure 4(a) shows the fitting for the daily increment based on the two-chain Fudan CCDC model. There are two peaks in the curves, suggesting possible new sources of transmission. Then we single out the ‘most optimized’ curve (the red line) and draw its area (in light blue) of ±25% fluctuation in Figure 4(b). We see that most of the data fall in this area, indicating the effectiveness of the model. Besides, the two chains are shown in green and blue dotted lines, respectively.

### 2.2 Results of the three-chain model

Now we consider the three-chain Fudan-CCDC model. Figure 5(a) and 6(a) show the epidemic evolution in Singapore based on the three-chain model. The legends are the same as in the previous context. We can see from Figure 5(a) that under the three-chain model, Singapore is expected to have zero increment of confirmed cases on May 4, and the total number of infections will be around 1900, if no other chains of transmission arise in the future.

**Figure 5:**
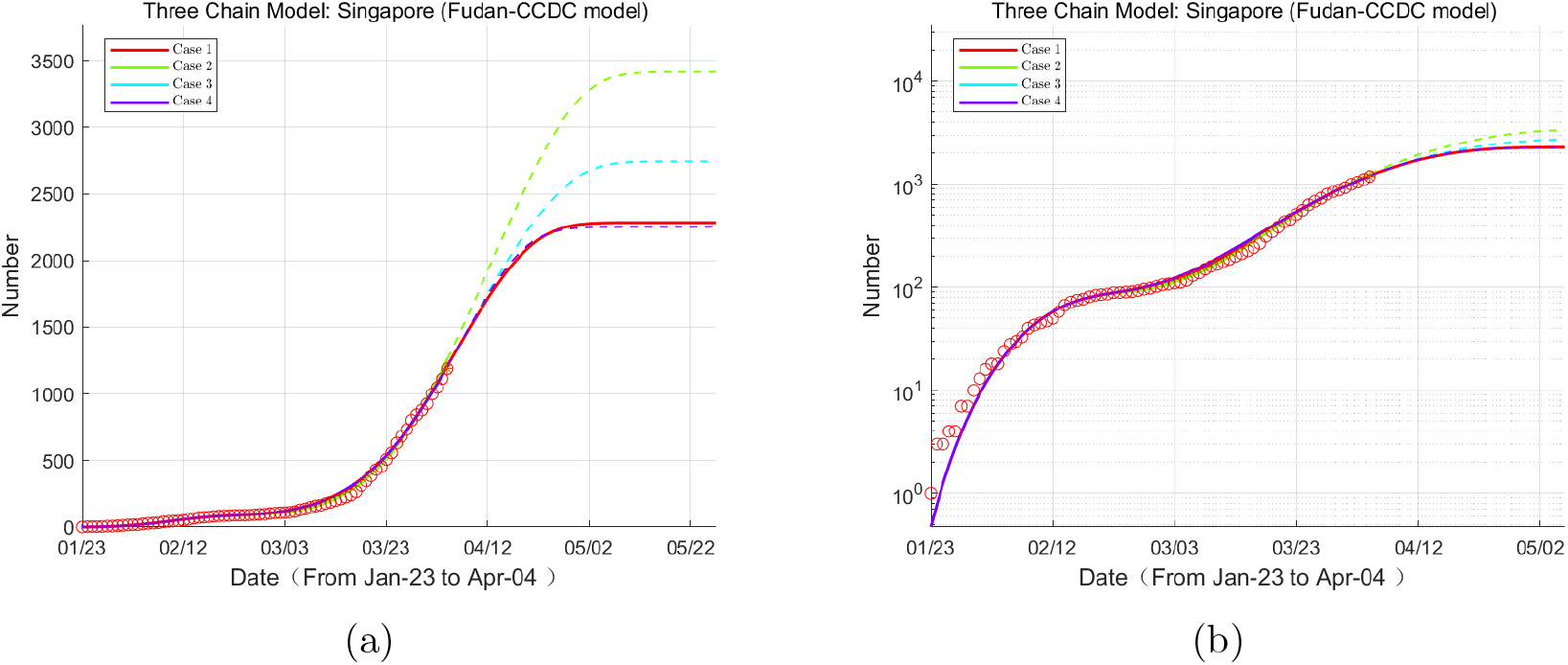
(a) Evolution of number of cumulative confirmed cases based on the three-chain Fudan-CCDC model. (b) The semi-log form of (a). The legends are the same as in Figure 3.

In addition, we find that the end date of COVID-19 based on the three-chain model is later than that based on the two-chain model, and the number of total infected is also significantly higher. This is because the time of zero increment will now arrive until all the transmission chains come to end.

### 2.3 Warning a possible new outbreak in Singapore

Figure 7 and Figure 8 show predictions of the cumulative and incremental confirmed cases in Singapore based on the multi-chain model, with data observed from Jan 23 to Apr12. The predictions of the two-chain model shows an uncontrollable trend of the epidemic. In the three-chain model, cumulative confirmed cases would become stable in late May, peaking at around 7200. These two models both pass on the information that Singapore might be faced with a very risky situation of rapidly increasing cases. Therefore, strong measures are urgently needed to contain the epidemic.

**Figure 6:**
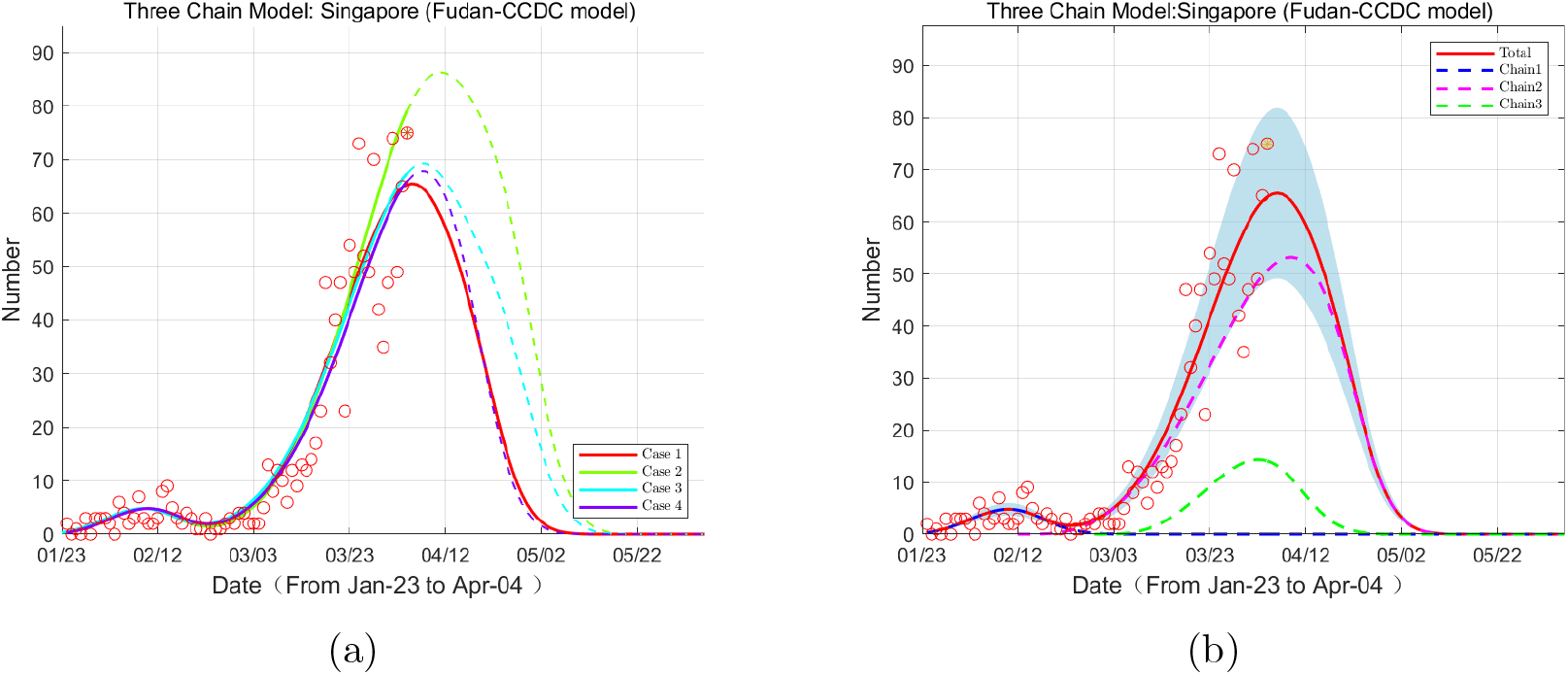
Evolution of the increment of confirmed cases based on the three-chain Fudan-CCDC model. The legends are the same as in Figure 4.

**Figure 7:**
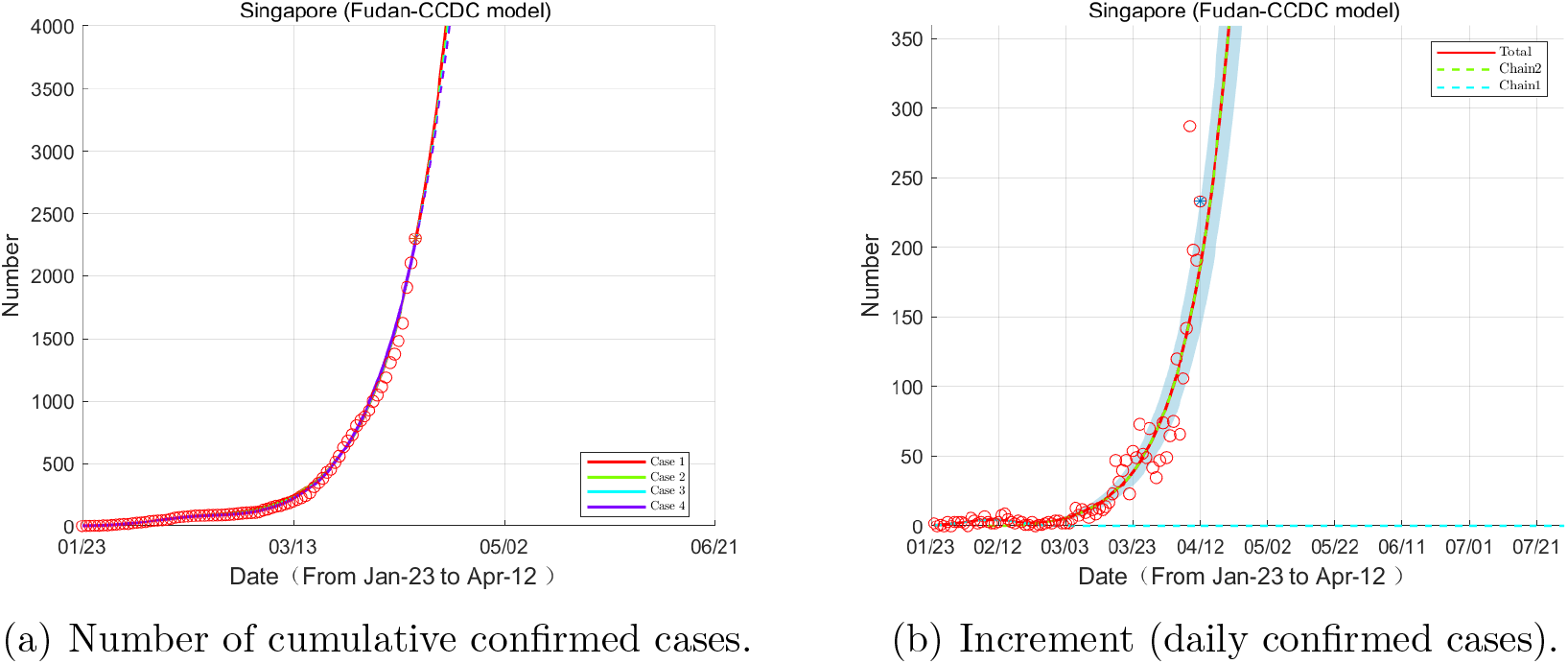
Evolution of COVID-19 based on the two-chain model, Jan 23 – Apr 12.

**Figure 8:**
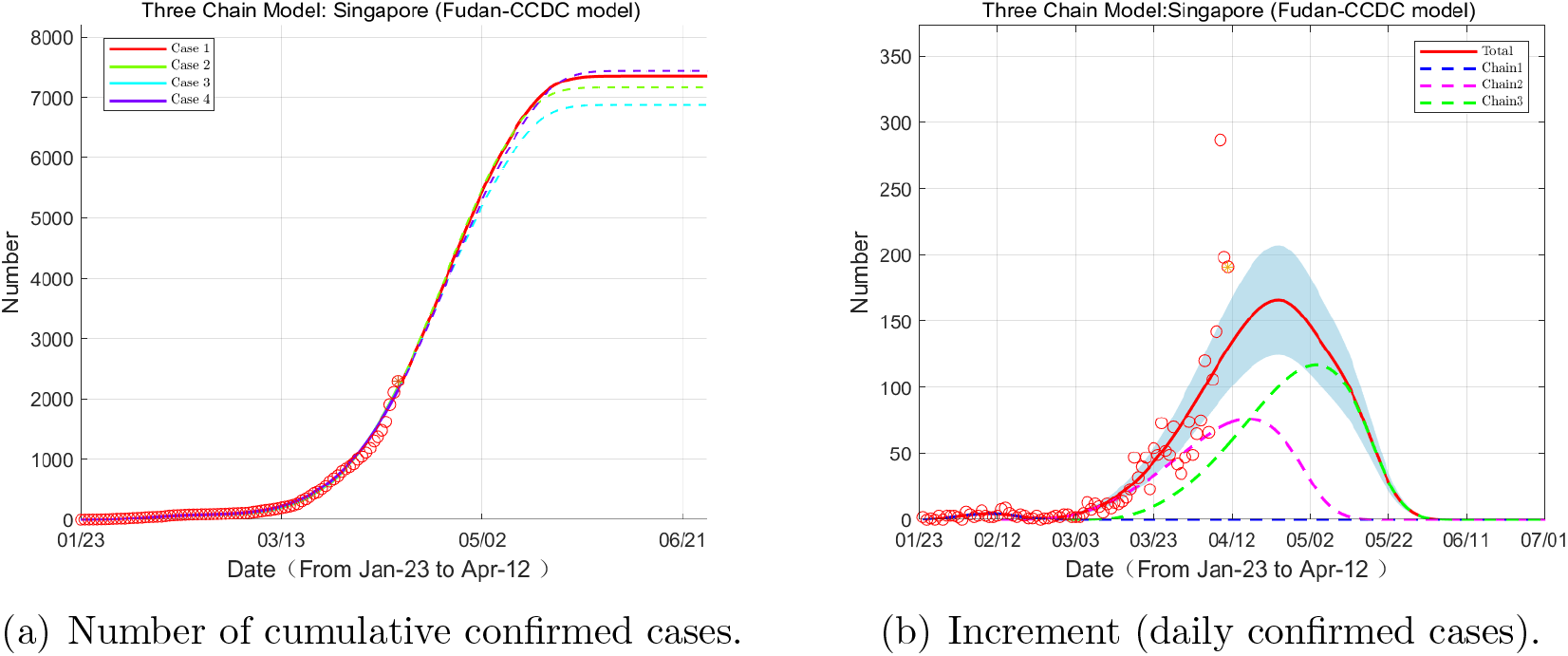
Evolution of COVID-19 based on the three-chain model, Jan 23 – Apr 12.

## 3 Discussion

### Advantages of the multi-chain Fudan-CCDC model

The multi-chain Fudan-CCDC model has given a better explanation of the epidemic evolution in Singapore and Iran, and perhaps other nations as well. Compared to single chain models, the multi-chain Fudan-CCDC model shows the following advantages: (1) It better fits the data in history; (2) By identifying different sets of parameters for different chains, it is able to simulate the multi-peaks in the daily increment data, which the single-chain models can hardly explain; (3) It illustrates the importance of controlling the imported cases. Since zero increment depends on when the last chain vanishes, it is difficult to completely end the epidemic unless all sources of transmission are detected and blocked.

### Detection of new chains

Now we revisit Figure 2 to discuss when to introduce new chains. In Figure 2(b), there is an obvious shallow pit around Feb 27 along the data trend, illustrating that the number of confirmed cases was about to flattened, but rose up again immediately. This shallow pit acts as a signal to consider new chains in the model, warning new sources of transmission. In fact, in Figure 2(a), a shallow pit has already occurred around Feb 12. This pit was not so obvious as the next one around Feb 27, and was likely to be treated as fluctuation of the data. Besides, more data are needed to form a new transmission chain. Therefore, carefully detecting and analyzing these shallow pits plays an important role in finding new chains.

### Additivity

One may find that the multi-chain model is just an addition of multiple single-chain models. In fact, the single-chain and multi-chain Fudan-CCDC models are both linear ones, so they enjoy the convenience of additivity. It is a major feature of our model, and it is not easy to get such good property in traditional epidemic models such as the SIR, SEIR, SEIJR models, which are all nonlinear. The property of additivity is friendly, as it allows us to construct new models of not only multiple chains, but also multiple districts, which might be applicable to Iran, the European Union, the United States and the whole world. We have already got some good simulation results for the epidemic in Iran, and more results on more countries will be reported in the near future.

In conclusion, the multi-chain Fudan-CCDC model is suitable for Singapore. It has made possible the early detection of imported infectors and super spreaders, and is able to suggest timely adjustment for epidemic control. Our results show that Singapore is currently in a more risky situation than before because of the new transmission chains brought by the imported infectors. As is mentioned in our previous paper [14], the Singapore government has always been making great efforts in trace-tracking and disease prevention, and there is much experience by other countries to learn from, so we believe that Singapore has the capacity to handle the crisis and eventually end it, as long as they keep alert on travellers and cut down every chain of transmission.

## 4 Materials and Methods

In this section, we introduce two models, the single-chain Fudan-CCDC model and the multi-chain Fudan-CCDC model respectively. The single-chain Fudan-CCDC model describes the evolution of COVID-19 based on the assumption that all the new cases originate from the initial source, i.e. there is only one chain of transmission. And the multi-chain Fudan-CCDC model assumes that due to new imported cases, new super spreaders, or the different transmission characteristics of different regions, there may be two or more single chains of transmission in the country.

### 4.1 The single-chain Fudan-CCDC model

As is mentioned in [10, 8, 9, 12, 14], our single-chain Fudan-CCDC model is as follows:

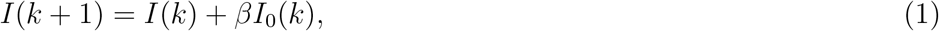

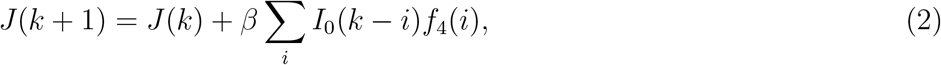

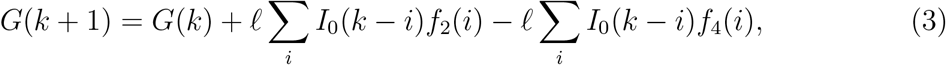

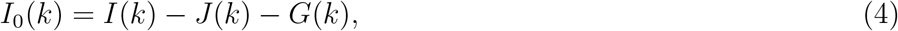

where *I*(*k*) and *J* (*k*) represent the cumulative infected people and the cumulative confirmed cases at day *k*, respectively, and *G*(*k*) is the instant (not cumulative) number of infected isolated not yet confirmed by the hospital. The infected ones are put into isolation once they show illness symptoms, and the newly confirmed should be removed from the isolated group. *I*_0_(*k*) is the number of people who are potentially infectious to healthy ones–they are infected actually but not in quarantine or hospitalization. *β* and *ℓ* represent the infection rate and the isolation rate respectively, which may be changed in different time periods. *f*_2_(*k*) and *f*_4_(*k*) are the transition probabilities from infection to illness onset, and from infection to hospitalization, respectively, which are reconstructed from one important paper [5] by CCDC. This time delay dynamic system is applicable to simulations of COVID-19 in the countries where community transmission exists, while the kernels like *f*_2_(*k*) and *f*_4_(*k*) might vary from countries to countries.

The model can be used to fit the reported numbers of the cumulative confirmed cases and predict the evolution of epidemic, and the details can be found in [10, 8, 9, 12, 14].

### 4.2 The multi-chain Fudan-CCDC model

In the multi-chain Fudan-CCDC model, the final epidemic transmission chain is the superposition of several single chains:

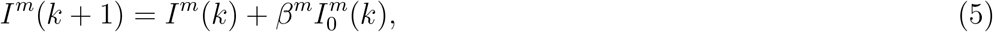

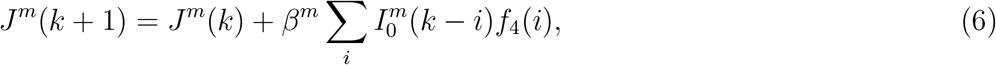

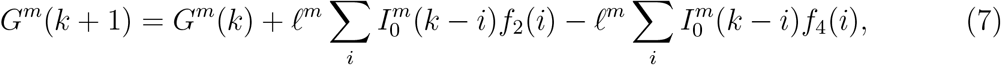

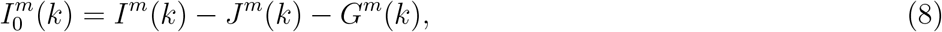

and we obtain the sum forms:

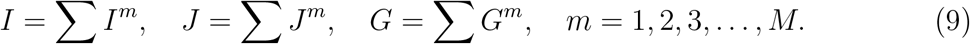

where *t*^*m*^ is the start time of the *m*-th source.

Specifically, we have applied the two-chain and the three-chain models to analysis the situations in Singapore. We suppose that there is a new chain when a sudden turn appears in the curve of reported confirmed cases. For both the two-chain model and the three-chain model, infection rate *β* and isolation rate *ℓ* of the fisrt chain are obtained by fitting data before a specific time node. The differences lie in assumptions and parameters to identified.

### 4.3 Optimization method for parameter identification

Parameter identification is an optimization process. There are two kinds of decision variables in this optimization, time nodes *t* and the model parameters. We suppose that more recent data have more importance and efficiency for us to predict the trend. So we established the objective function as follows:

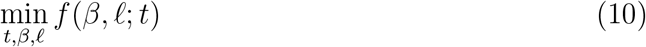

where 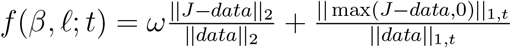.

Note that the first term is to minimize the difference betweem values of data and simulations, i.e. the empiral risk and the second term is to minimize the structural risk. *ω* is the weight of the penalty term, and *t* contains only *t*^2^ in the two-chain model, and (*t*^2^, *t*^3^) in the three-chain model.

#### Time nodes

Since public data are discrete along time and time nodes are dates, the grid searching method could be used to obtain the minimum value of the objective function. As characteristics of the first chain is known, so we only need to do grid searching of other chains.

#### Model parameters

The parameter optimization is solved by a constrained optimization problem solver.

Therefore, the whole process of optimization can be summarized as the following three steps: determine all possible time nodes; calculate the minimum of objective function for cases of different time nodes; obtain the optimal time nodes and model parameters.

### 4.4 Values of the parameters

In Figure 4, we can clearly see the rise of another chain, and the lifting of the second chain is more independent of the first chain, i.e., the second chain emerges when the first chain almost subsides. The parameters of the first chain, *β*^1^ and *ℓ*^1^, are obtained by fitting data before Feb 17, considering that the output curve remain stable for a long period. It is noticed that parameters identified on later date are more likely to be affected by the new source. Assuming that *ℓ* in one region is relatively stable, we set fixed value for isolation rate, i.e. *ℓ*_1_ = *ℓ*_0_.

The values of the parameters in the two-chain model and the three-chain model are listed in Table 1. From this table, we think that the three-chain model is more reasonable for the case of Singapore, since the important parameter *β* is close in the three-chain model, and we mention that *β* is essential to the estimation of the basic reproductive number [9].

**Table 1:** Values of the parameters identified by the multi-chain models.

|  | the two-chain model |  |  | the three-chain model |  |  |
| --- | --- | --- | --- | --- | --- | --- |
| | start date | $\beta$ | $\ell$ | start date | $\beta$ | $\ell$ |
| chain 1 | Jan15 | 0.33704 | 0.50507 | Jan 15 | 0.33704 | 0.50507 |
| chain 2 | Feb 12 | 0.45546 | 0.66223 | Feb 13 | 0.37684 | 0.50507 |
| chain 3 | - | - | - | Feb 21 | 0.36985 | 0.50507 |

## Data Availability

All the data can be accessed publicly

https://www.who.int

## Acknowledgments

We are very grateful to the efforts of Cheng’s group members and the supports by School of Mathematical Sciences, Fudan University and School of Mathematics, Shanghai University of Finance and Economics. We thank Zhaojun Bai at UC Davis, Xiaoming Wang at South University of Science and Technology of China, Qiang Du at Columbia University, Long Chen at UC Irvine, Cheng Wang at University of Massachusetts at Dartmouth and Xingjie (Helen) Li at University of North Carolina at Charlotte. Wenbin Chen also thanks Wei Ge and Rongmin Li at Fudan University, Zhihua Shen at Fusion Fin Trade, Allen at Wind, Ning Liu at Winning Health Technology Group Company (winning.com.cn), Xinkang Cao and Jinwu Zhuo at Mathworks, and Weili Zhang. All of us thank our families’ supports.

## Ethics approval and consent to participate

The ethical approval or individual consent was not applicable.

## Authors contributions

The algorithms are implemented by Hanshuang Pan, which are based on the single-chain model implemented by Nian Shao, and designed by Wenbin Chen. All authors conceived the study, carried out the analysis, discussed the results, drafted the first manuscript, critically read and revised the manuscript, and gave final approval for publication.

## Data and materials availability

The data employed in this paper are acquired from WIND (like Bloomberg), and the situation reports of the World Health Organization (url: https://www.who.int). All the data can be accessed publicly. No other data are used in this paper.

